# An in-vivo approach to quantify intra-MRI head motion tracking accuracy: comparison of markerless optical tracking versus fat-navigators

**DOI:** 10.1101/2025.04.23.25326185

**Authors:** Zakaria Zariry, Franck Lamberton, Robert Frost, Thomas Gaass, Thomas Troalen, Holly Rayson, Jakob M. Slipsager, Nathalie Richard, Andre van der Kouwe, James Bonaiuto, Bassem Hiba

## Abstract

**Purpose:** Head-motion tracking and correction remains a key area of research in MRI, but the lack of rigorous and standardized evaluation approaches hinders their optimization and comparison. We introduce an in-vivo framework for assessing the accuracy of intra-MRI head motion tracking, and demonstrates its effectiveness by comparing two methods based on a markerless optical system (MOS) and a fat signal navigator (FatNav).

**Methods:** Six participants underwent 3T brain MRI using a T1-weighted (T1w) pulse-sequence with a fat-navigator module. Participants performed head-rotations of 2° or 4°, each visually guided by MOS feedback around a single primary axis (X or Z). MOS and FatNav estimations were evaluated against rigid-registration of T1w-images, as gold-standard, across seven different head positions.

**Results:** The proposed approach revealed that MOS outperforms FatNav in estimating translation and large head rotations (2-4°), while FatNav shows better accuracy for subtle rotations. Image quality assessments following correction for three head rotations (rightward, upward, and leftward) confirmed that MOS outperformed FatNav in restoring image fidelity, as evidenced by the higher Structural Similarity Index, Peak Signal-to-Noise Ratio, and Focus Measure. Unlike the traditional image quality-based comparisons, the proposed framework demonstrated sensitivity to subtle improvements in FatNav performance, achieved by applying a neck mask to the fat-navigator images.

**Conclusion:** The proposed framework enabled a precise in-vivo evaluation and comparison of MOS and FatNav for head-motions estimation. It was sufficiently sensitive to reveal a slight improvement in FatNav performance when neck was masked in fat-navigator images. In parallel, the conventional image quality-based approach confirmed the superior performance of MOS in restoring T1W image quality, though it did not capture the improvement achieved by FatNav with neck-masking. Together, these two complementary approaches provide a comprehensive assessment of both head-motions estimation and correction in MRI.

## 1. Introduction

Motion artifacts represent a prevalent source of degradation in magnetic resonance imaging (MRI). When left uncorrected, they can obscure subtle pathological changes and potentially lead to the misinterpretation of clinical images^1,2^. Furthermore, these artifacts incur substantial, undesirable costs for hospitals^3^. Andre et al. estimated that repeat sequences account for 19.8% of all MRI examination performed on inpatient and/or emergency department patients^4^. This issue is particularly critical for high-resolution brain MRI, which has gained renewed interest with the advent of 7T scanners^5^, and for imaging certain populations such as children^6^.

A multitude of approaches have been proposed to mitigate head-motion artifacts during MRI acquisitions. One class of techniques relies on so-called “navigators”. In brief, short acquisitions, timed before or after the echo-train at each repetition time (TR) of the imaging sequence, generate a series of low-resolution three-dimensional images of the head. These images are then registered, either prospectively or retrospectively, to estimate and compensate for head-motion at each TR. The fat navigator (FatNav) strategy employs spectrally selective RF excitation to acquire subcutaneous fat images, which serve as a head-motion navigator^7,8^.

An alternative strategy involves external devices designed to monitor the head’s position and orientation relative to the scanner. For example, electroencephalography sensors^9^, electromagnetic sensors^10^ and optical tracking cameras^11,12^ have been proposed to track and correct rigid head-movements during MRI. In parallel, advanced image acquisition and reconstruction techniques^13^, often data-driven and increasingly powered by artificial intelligence^14,15^, are also under active investigation.

Given the proliferation of head-motion correction methods in MRI, there is an urgent need for rigorous evaluation strategies that not only allow radiologists and neuroimaging researchers to appreciate the image quality benefits of each technique but also guide developers in refining their approaches. Several research groups have utilized head-phantoms affixed to mechanical positioning systems capable of precise rotational and translational displacements to establish the prescribed motion parameters as a gold-standard reference for motion tracking validation. Errors in phantom motion tracking are then directly quantified by comparing the estimated rotations/translations values against the gold-standard reference values^16,17^. However, the complex anatomy and composition of the human head, and its actual movements governed by complex biomechanical constraints, are not easily replicable using phantoms. Moreover, external motion sensors like optical cameras may produce different estimates for an identical movement when performed by a human head compared to a phantom, due to inherent differences in appearance and texture. Finally, Gumus et al. found that head motion tracking errors measured in-vivo exceeded those detected in offline tests^18^.

In the absence of an in-vivo gold-standard for evaluating the accuracy of motion estimation, indirect assessment methods, typically involving the comparison of motion-corrected images with presumed motion-free reference images^19,20^, are commonly employed in-vivo. However, such approaches, which rely on image quality metrics, brain morphometric fidelity^6,16^, or the preservation of clinically relevant features^21^, may be subject to biases stemming from the initial quality of the reference motionless images and from the limited sensitivity or relevance of the employed image quality metrics^20^. To date, only one study, by Gumus et al., has established an in-vivo gold standard for head-motion tracking^18^. This approach used SPM-registration of brain 3D T1-weighted (T1w) images acquired in various head positions relative to a reference pose^18^. The head motion estimates from the tracking method were then validated against rotations and displacements computed through 3D T1w-images rigid registration.

Despite ongoing advances in head motion tracking and correction techniques in MRI, the absence of robust and standardized in-vivo evaluation protocols remains thus a critical bottleneck, hindering not only the precise optimization and objective comparison of these methods but also their broader application beyond anatomical purposes. To address this challenge, the present study proposes an in-vivo evaluation framework for head motion tracking and correction, incorporating a FreeSurfer registration-based gold-standard and an intra-MRI visual instruction system designed to guide and standardize head movements across multiple subjects. We employed this methodology to evaluate and compare the performance of two techniques, one based on a markerless optical-system^16^ (MOS) and the other on the 3D FatNav strategy^8^, in six healthy subjects.

We further challenged the proposed approach for assessing the purported improvement in FatNav performance resulting from the application of a neck mask to fat navigation images^22^, and investigated the validity of using FreeSurfer-based registration as a gold-standard for tracking head motion during MRI.

## 2. Materials, methods, and participants

### 2.1. MRI materials and participants

MRI acquisitions were conducted on a MAGNETOM Prisma 3 T scanner (Siemens Healthcare, Erlangen, Germany) equipped with a 64-channel receive coil. Six healthy volunteers (31.5±6.2 years) were recruited in accordance with the Declaration of Helsinki, and all participants gave written informed consent which was approved by the regional ethics committee for human research (CPP Sud Mediterranee V-2021-A00042–39).

A magnetization-prepared rapid gradient echo (MP-RAGE) pulse sequence, incorporating a Fat-navigator module^8^ following each TR, was employed for sagittal T1w-image acquisition. Simultaneously with the MP-RAGE acquisition, head movement parameters were recorded every ∼33 ms using the markerless optical device Tracoline (TCL) in combination with the TracSuite v3.3 software^23^ (TracInnovations, Ballerup, Denmark). The TCL probe was positioned adjacent to the head-coil to capture part of the facial surface through the coil’s aperture in front of the right eye (Supplementary data, figure 1). This system utilizes active stereo vision in conjunction with a structured infrared light approach to acquire three-dimensional point clouds of the subject’s face at a rate of approximately 30 frames per second. These point clouds are subsequently co-registered with a reference model to compute rigid body pose estimates. A TCL/MRI-scanner cross-calibration procedure^24^ to determine the transformation between optical and MRI coordinates was performed before each MRI session.

### 2.2. Motion-correction and image reconstruction

After acquisition, the Fourier-space data were transferred to a dedicated server for motion correction and image reconstruction. The RetroMocoBox toolbox^25^ was then used to reconstruct the 3D fat-navigator images, before rigidly realigning them to estimate the head-movement parameters at each TR, relative to the first-TR. Two sets of motion-parameters (three rotations and three translations) were computed for each MP-RAGE-image based on: 1) the original 3D fat-navigator images; 2) 3D-fat-navigator images after neck-masking^22^. A custom Python pipeline was developed to apply motion transformations, estimated using either MOS or FatNav, to each Fourier-space line after completing the missing data with the GRAPPA algorithm^26^. The final T1w-image was reconstructed using a non-uniform inverse Fourier transform.

### 2.3. Experimental setup and head motion control

To maintain uniform and reproducible head movements among participants, custom Python-based software was developed to provide visual instructions and feedback within the MRI environment. The 2D visual instruction animation was projected onto a screen at the back of the MRI tunnel via an external projector positioned outside the MRI room, facing a waveguide. Participants viewed the screen through a mirror mounted atop the 64-channel coil. The animation featured a white background with a red square that dynamically reflected the subject’s head position, as measured in real time by the MOS system, and a square with blue frame indicating the target position (Figure 1). The participant is instructed to align the red square to the blue one by adjusting their head rotations. The red square’s lateral displacement corresponded to rotations around the Z-axis, while its vertical movement was linked to rotations around the X-axis. Additionally, a thin black border surrounding the red square rotated according to the Y-axis rotation. A text message appeared five seconds before each movement to alert participants and prevent abrupt or forceful motions. To maintain participant comfort and prevent excessive instruction overload, we restricted visual guidance to head rotations only.

**Figure 1.**
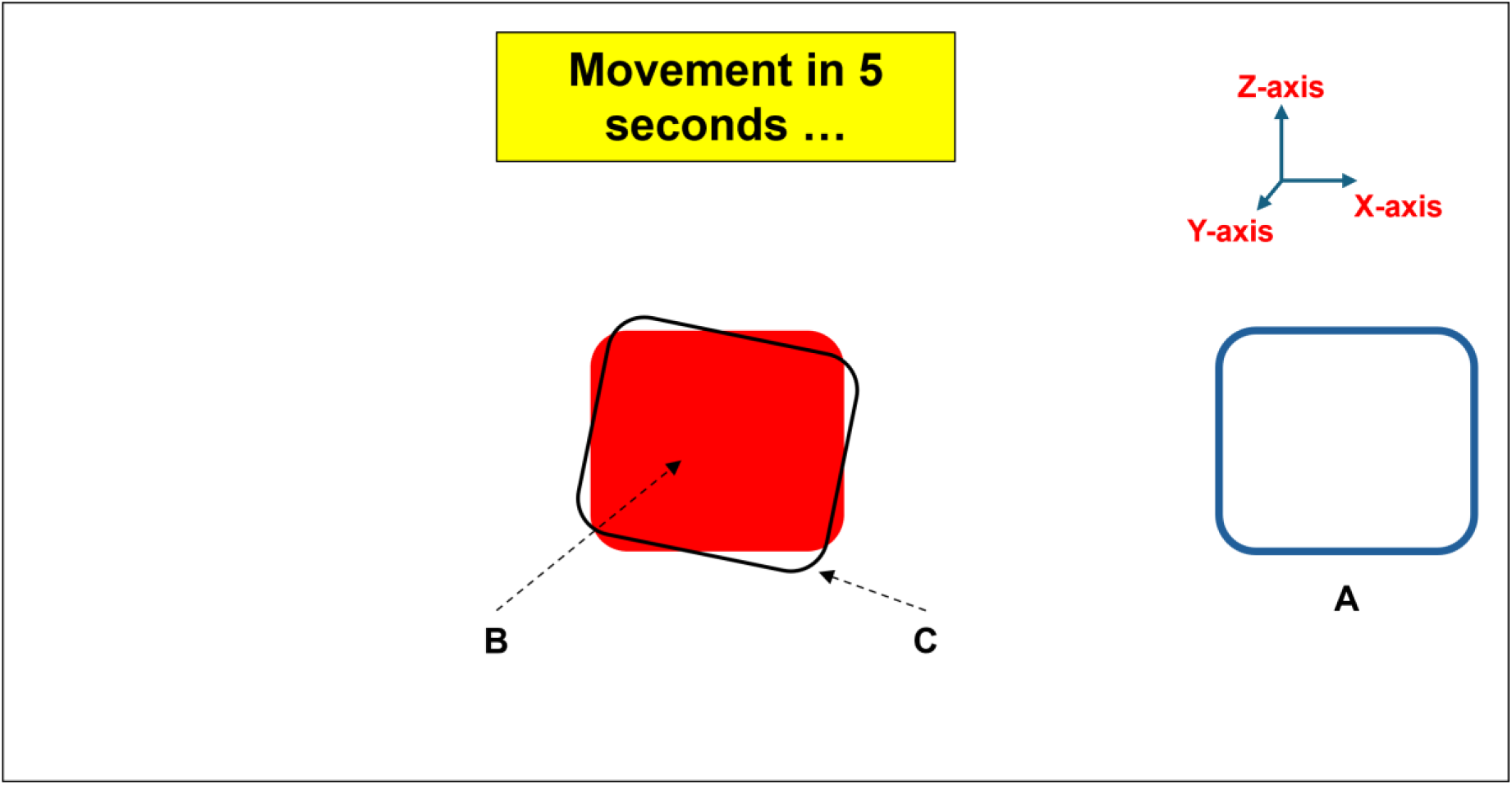
Visual instructions projected within the MRI to guide head rotations. The vertical and horizontal displacements of Square-A and Square-B from the isocenter correspond to the head rotations around the X- and Z-axes, respectively. Square-A represents the real-time head rotation as measured by the motion tracking system (MOS), whereas Square-B indicates the target head position. Square-C rotations, relative to Square-B, provide real-time feedback on head rotation around the Y-axis. In the present configuration, participants are instructed to perform a 4° rightward head rotation until Square-B translated horizontally to align with Square-A.

Participants performed head rotations rightward (yaw^-^), upward (pitch^+^), and leftward (yaw^+^) of 2° or 4° around a single primary axis (X or Z). Associated minor head rotations were observed along the secondary axes of each rotation (Primary rotation around the X-axis iq accompanied by minor secondary rotations around the Y- and Z-axes, while primary rotation around the Z-axis involved minor secondary rotations around the X- and Y-axes). Similarly, head rotations induce translations in all three spatial directions (Supplementary data, figure 2).

### 2.4. MRI experiments

#### 2.4.1. Experiment-1: Head-motion estimation accuracy of MOS vs. FatNav

This experiment aimed to estimate movement parameters in degrees and millimeters using the MOS and FatNav methods for various stereotypical head poses relative to a reference position. The T1w- image acquired at each head-pose was registered to the reference-position image using FreeSurfer’s *mri_robust_register* tool^27^ This procedure yields a transformation matrix describing an ideal rigid alignment between the two head positions, which is considered the gold-standard. Finally, the rotational and translational errors for each head-position, for both MOS and FatNav, were computed by comparing their measurements against the gold-standard estimation, as explained in supplementary data figure 3. Since the MP-RAGE pulse sequence follows a sequential phase encoding scheme, the FatNav and MOS head locations for each T1w-image were determined based on the fat-navigator location closest to the Fourier-space center sampling and the average MOS locations recorded during the five seconds nearest to this sampling.

Each participant underwent seven separate MP-RAGE scans with an isotropic spatial resolution of 1mm and acquisition parameters optimized to limit its duration to 4 minutes 25 seconds (Table 1). Before each T1w-scan, participants adjusted their head to a target position displayed on the screen and remained still. The first scan, with the head straight and centered, served as a reference. Before the next three scans, participants performed yaw^-^, pitch^+^ and yaw^+^ rotations of 2° relative to the reference position. These rotations were then repeated with a 4° amplitude for the final three scans.

**Table 1.** Parameters of sagittal T1-weighted MP-RAGE pulse sequence and its FatNav module.

| Acquisition parameter | Experiment-1 | Experiment-2 |
| --- | --- | --- |
| Isotropic resolution (mm) | 1 | 1 |
| Field-of-view (mm<sup>3</sup>) | 256×256×192 | 256×256×192 |
| Inversion time (ms) | 900 | 1070 |
| Echo time (ms) | 3.3 | 3.3 |
| Repetition time (ms) | 2180 | 2500 |
| Flip angle (°) | 8 | 8 |
| Bandwidth (Hz/Pixel) | 250 | 240 |
| Parallel imaging: Acceleration factor | 3×1 | 2×1 |
| Parallel imaging: Nb. of ref. lines | 40 | 32 |
| Turbo factor | 208 | 192 |
| Nb. of 3D fat-navigator images | 112 | 143 |
| FatNav - resolution (mm) | 4 |  |
| FatNav - TE/TR (ms) | 1.43/3.4 |  |
| FatNav - Flip angle (°) | 7 |  |
| FatNav – Acceleration factor | 4×4 |  |
| FatNav - Bandwidth (Hz/Pixel) | 1950 |  |
| Total acquisition time | 4 min 9 sec | 6 min 2 sec |

**Table 2.** Mean number and ratio of brain voxels with distortion vector norms greater than 0.5 mm in T1- weighted images from Experiment-1. The distortion field was derived from elastic and affine registrations between each T1w-image and its corresponding reference.

| Rotation | N. of brain voxels | N. of voxels with distortion norm > 0.5 mm | Ratio of voxels with distortion norm > 0.5 mm |
| --- | --- | --- | --- |
| Yaw<sup>+</sup> 2° | 1439566 | 2014 | 0.14 % |
| Yaw<sup>+</sup> 4° | 1439566 | 8245 | 0.57 % |
| Yaw<sup>-</sup> 2° | 1439566 | 5035 | 0.35 % |
| Yaw<sup>-</sup> 4° | 1439566 | 24442 | 1.7 % |
| Pitch<sup>+</sup> 2° | 1439566 | 36235 | 2.5 % |
| Pitch<sup>+</sup> 4° | 1439566 | 41578 | 2.89 % |

To account for T1w-image distortion variations related to head positioning, rigid, affine, and non-linear registrations were performed using ANTs^28^ between each T1w-image and its corresponding reference image. The distortion-corrected image was then transformed back into its native space using the inverse of the rigid transformation. Finally, the FreeSurfer rigid registration was applied to obtain a distortion-corrected gold-standard for head motion estimation.

#### 2.4.2. Experiment-2: Image quality recovery by MOS vs. FatNav

For each participant, three additional T1w-images (6 minutes each) were acquired, with settings adjusted for optimal gray to white-matter contrast (Table 1). The participant was instructed to keep their head still at the reference position for the first image. During the second acquisition, the participant was visually instructed to center their head before executing three head rotations of 2° relative to the reference position: yaw^-^; pitch^+^; and yaw^+^, at 1 minute 20 seconds; 2 minute 30 seconds; and 4 minutes respectively after the scan began. The third acquisition followed the same head rotations with amplitude increased to 4°. T1w-images were each reconstructed three times using the retrospective motion-correction pipeline with motion estimates derived from MOS, FatNav, and neck- masked FatNav respectively. The reference T1w-image was corrected for any unintended head-movement occurring during its acquisition using MOS estimates.

Brain extraction was then carried out using the BET^29^ tool. Each T1w-image was aligned with the reference image using *mri_robust_register* to facilitate quantitative quality assessments based on three metrics: Structural Similarity Index (SSIM)^30^, Peak Signal-to-Noise Ratio (PSNR), and Focus Measure (Variance of Laplacian)^31^. Finally, a visual quality evaluation of motion-corrected images was performed by three independent blinded experts who evaluated three key criteria: sharpness, noise, and ghosting artifacts, using a scoring system from 1 (poor) to 5 (excellent).

### 2.5. Statistical analysis

Pairwise statistical tests were performed on the estimation errors. To evaluate performance differences between MOS and FatNav, and between the two FatNav variants, a linear mixed-effects model was employed, including tracking method as a fixed effect, and subject-specific intercepts as random effects, treating different angles (2° and 4°) and directions (yaw and pitch) as repeated measures. These analyses were conducted using R (version 4.4.1)^32^ and *lme4* (version 1.1.36)^33^. Fixed effects were assessed using Type II Wald F tests with Kenward-Roger estimated degrees of freedom (*car* version 3.1.3)^34^. Pairwise Tukey-corrected follow-up tests were run using estimated marginal means from the *emmeans* package (version 1.10.6)^35^. A significance threshold of p<0.05 was adopted. Where applicable, effect sizes (standardized mean difference, Cohen’s *d*) were calculated to quantify the magnitude of the observed differences.

## 3. Results

### 3.1. Head motion reproducibility

The head-motion control strategy demonstrated high reproducibility across participants. For instructed 2° rotations, the gold-standard values were 2.09±0.39° for yaw^−^, -1.88±0.43° for pitch^+^, and - 1.98°±0.05° for yaw^+^. Similarly, for a 4° target head-rotation, the gold-standard values measured were 4.13±0.33°, -4.01±0.31°, and -4.08±0.22° in the respective directions. In addition to the expected rotations around the primary axes (Z for yaw and X for pitch), associated translations of the head and minor rotations around the secondary axes (X and Y for yaw and Y and Z for pitch) were also observed (additional data, Table 1).

### 3.2. Head-motion estimation accuracy: MOS vs. FatNav

A global analysis of Experiment-1 estimation errors covering the 108 rotation (6 subjects × 6 head positions × 3 axes) and the 108 translation indicates that both MOS and FatNav with neck-masking provided head movement estimates closely approximating the gold-standard (rotational and translational mean errors: 0.28 ± 0.25° and 0.33 ± 0.31 mm for MOS, and 0.28 ± 0.25° and 0.65 ± 0.68 mm for FatNav, respectively). While both methods performed similarly in estimating head rotations, MOS provided more accurate estimates of translational shifts (p<0.001. Figure 2).

**Figure 2.**
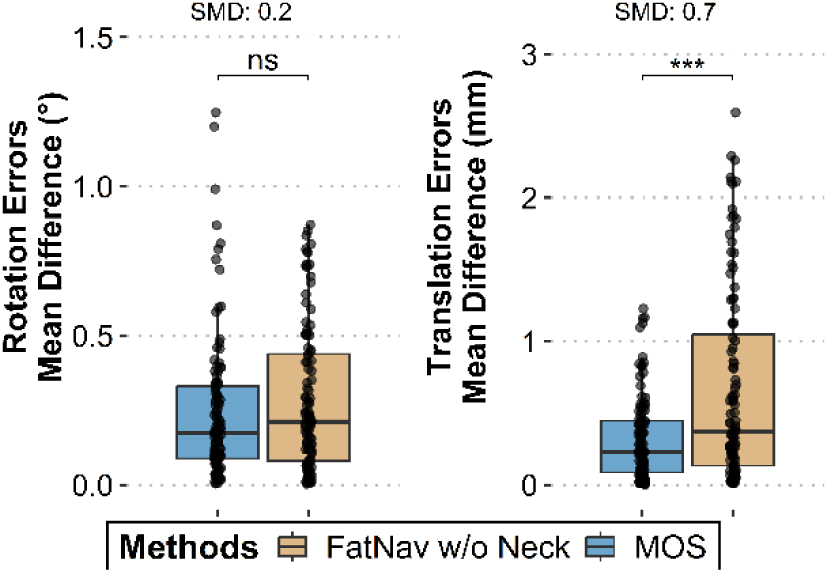
Global comparison of head motion rotation and translation estimation accuracy between MOS and FatNav with neck-mask.

For a more granular evaluation, performance differences between MOS and FatNav were independently assessed for yaw^-^, pitch^+^, and yaw^+^ rotations, with a separate analysis of large-amplitude rotations (2° and 4°) around the primary axes on one side, and smaller-amplitude rotations around the secondary axes on the other side. Figure 3 and supplementary data table 2 show results from Experiment-1 demonstrating that the mean errors for primary rotations were 0.21±0.23° for MOS and 0.50±0.22° for FatNav (8.4±11.6 and 17.5±8.1% respectively). MOS showed mean errors of 0.27±0.25° for secondary rotations and 0.32±0.29 mm for translations, whereas FatNav had mean errors of 0.17±0.17° and 0.66±0.68 mm, respectively (Supplementary data, table 2).

**Figure 3.**
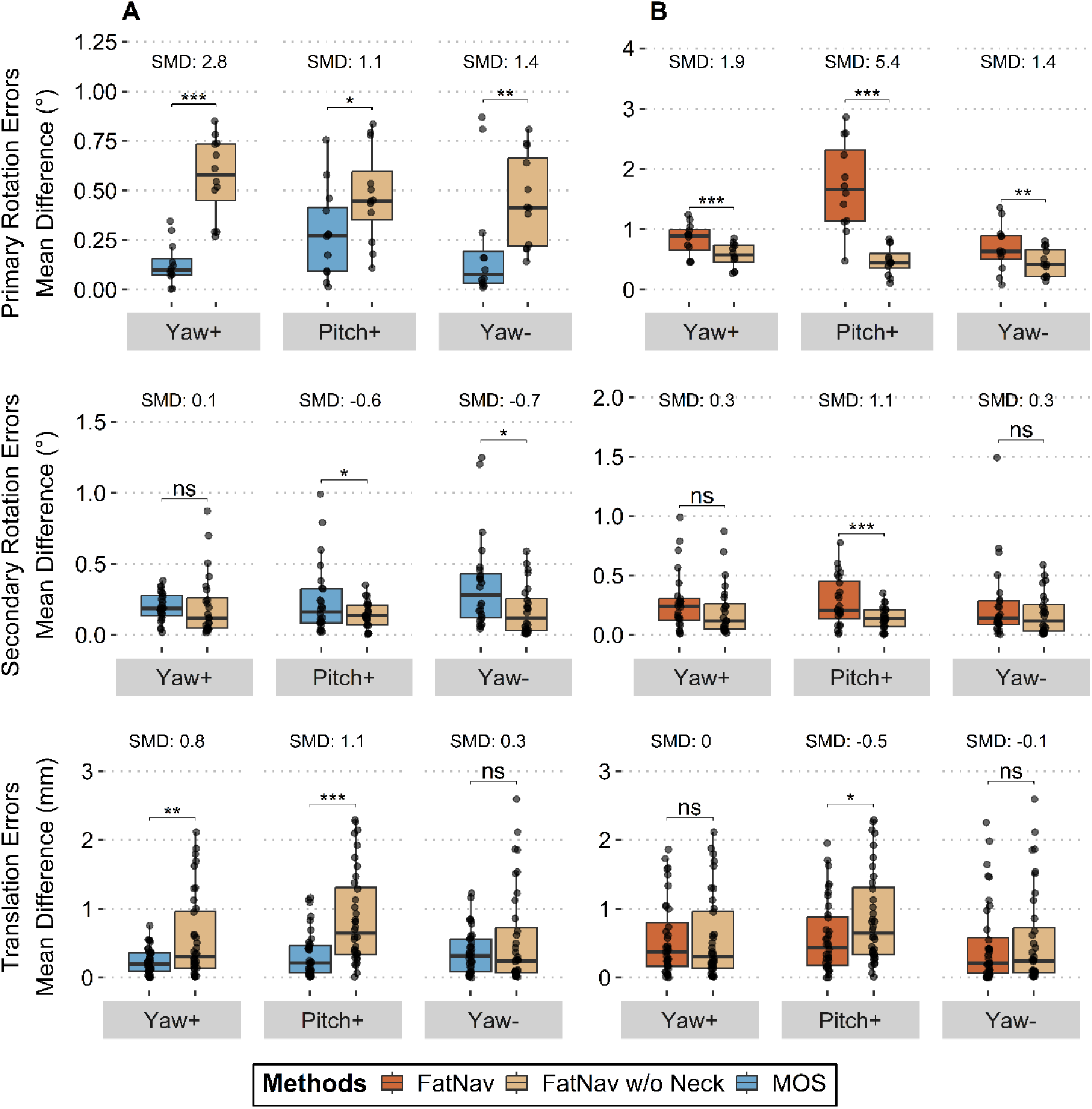
Accuracy of head-motion estimation for yaw^-^, pitch^+^, and yaw^+^ head movements, with a separate analysis of large-amplitude rotations (2° and 4°) around the primary rotational axes on one side, and smaller-amplitude rotations around the secondary rotational axes on the other side. A) MOS vs. FatNav wwith neck-mask and B) FatNav with and without neck-mask. SMD: Standardized mean difference.

Mixed model ANOVA (α=0.05) results confirmed that MOS reduced mean primary rotation errors by 38% for pitch^+^ (p=0.023), 77% for yaw^+^ (p<0.001), and 53% for yaw^−^ (p=0.004) compared to FatNav, demonstrating significantly better performance. MOS also demonstrated a superior performance on translation estimation, reducing mean errors of yaw^+^ by 59% (p=0.002) and of pitch^+^ by 60% (p<0.001), compared to FatNav.

In contrast, FatNav exhibited better accuracy than MOS for small rotations around secondary axes during yaw^-^ and pitch^+^, reducing mean errors by 50% (p=0.138) and 42% (p=0.039) respectively (Figure 3-A and supplementary data table 2). Interestingly, MOS’s ability to estimate yaw^+^ rotation was significantly more accurate than for yaw^-^ (p<0.05). Comparison of the two FatNav variants reveals that neck-masking significantly reduced the mean errors of primary and secondary pitch^+^ rotations by 48% and 32%, respectively (Figure 3-B).

To evaluate distortion in T1w images and its impact on gold-standard values, we computed the proportion of brain voxels with a displacement norm greater than half the voxel size (0.5 mm) from elastic and affine registration fields. Table 3 confirms mild T1w-image distortions that vary with the orientation and magnitude of head displacement relative to the reference position. The impact of this distortion is minimal, as head-motion estimates from both distortion-corrected and uncorrected gold standards are highly correlated (Figure 4). Furthermore, comparisons between MOS and FatNav remain largely unaffected when using distortion-corrected gold-standard values (Supplementary data, figure 4).

**Figure 4:**
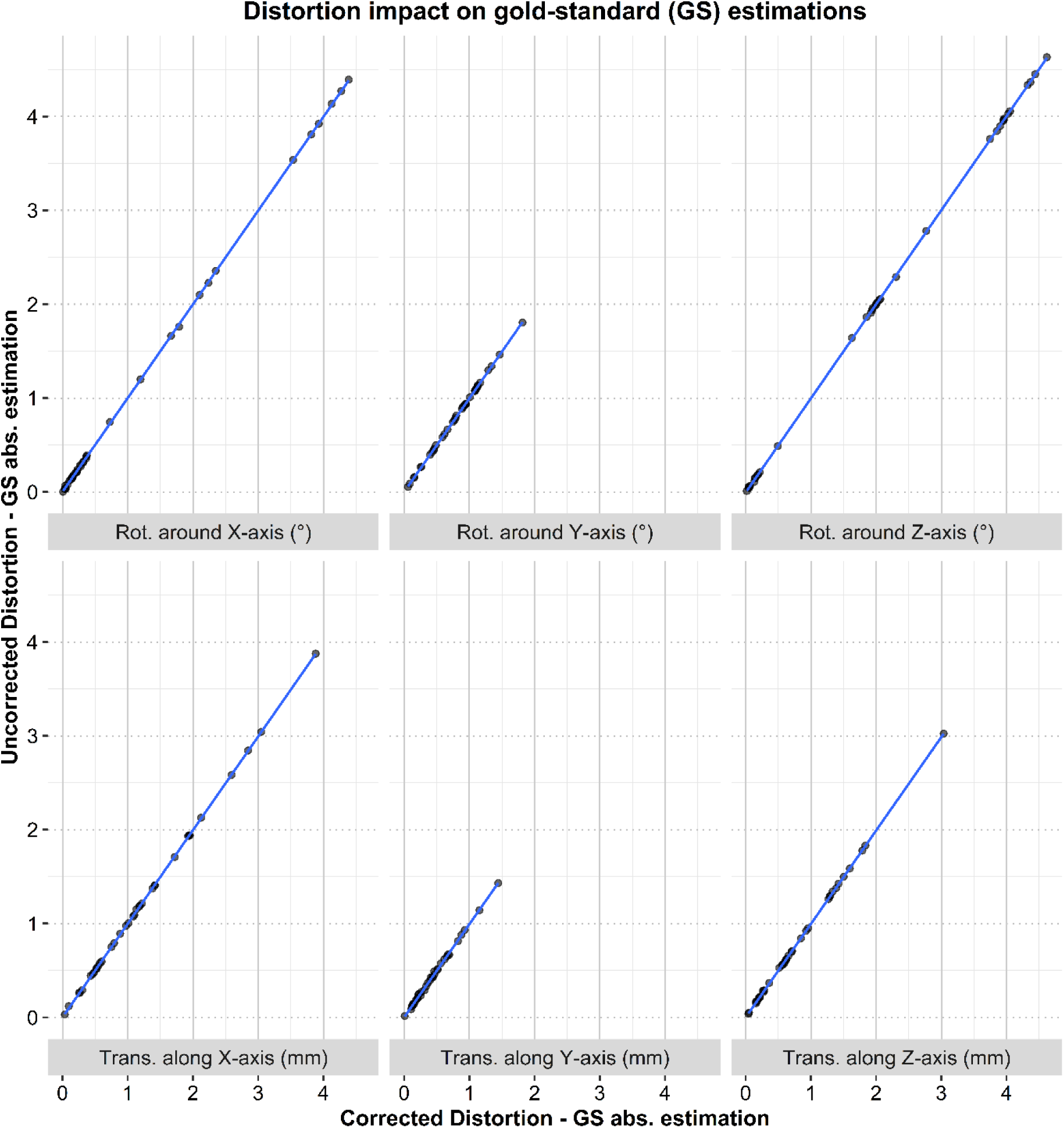
Comparison of head motion parameters of Experiment-1 derived from the rigid registration-based gold-standard and from the gold-standard with distortion-corrected T1w-images.

### 3.3. Image quality recovery by MOS vs. FatNav corrections

Figure 5 illustrates the impact of 2° head rotations on image quality in Experiment-2, alongside the retrospective motion correction capabilities of MOS and neck-masked FatNav. For reference, the figure also displays head rotations and translations along the X, Y, and Z axes, highlighting the discrepancies between MOS and FatNav measurements at each time point during a 6-minute T1w- acquisition.

**Figure 5.**
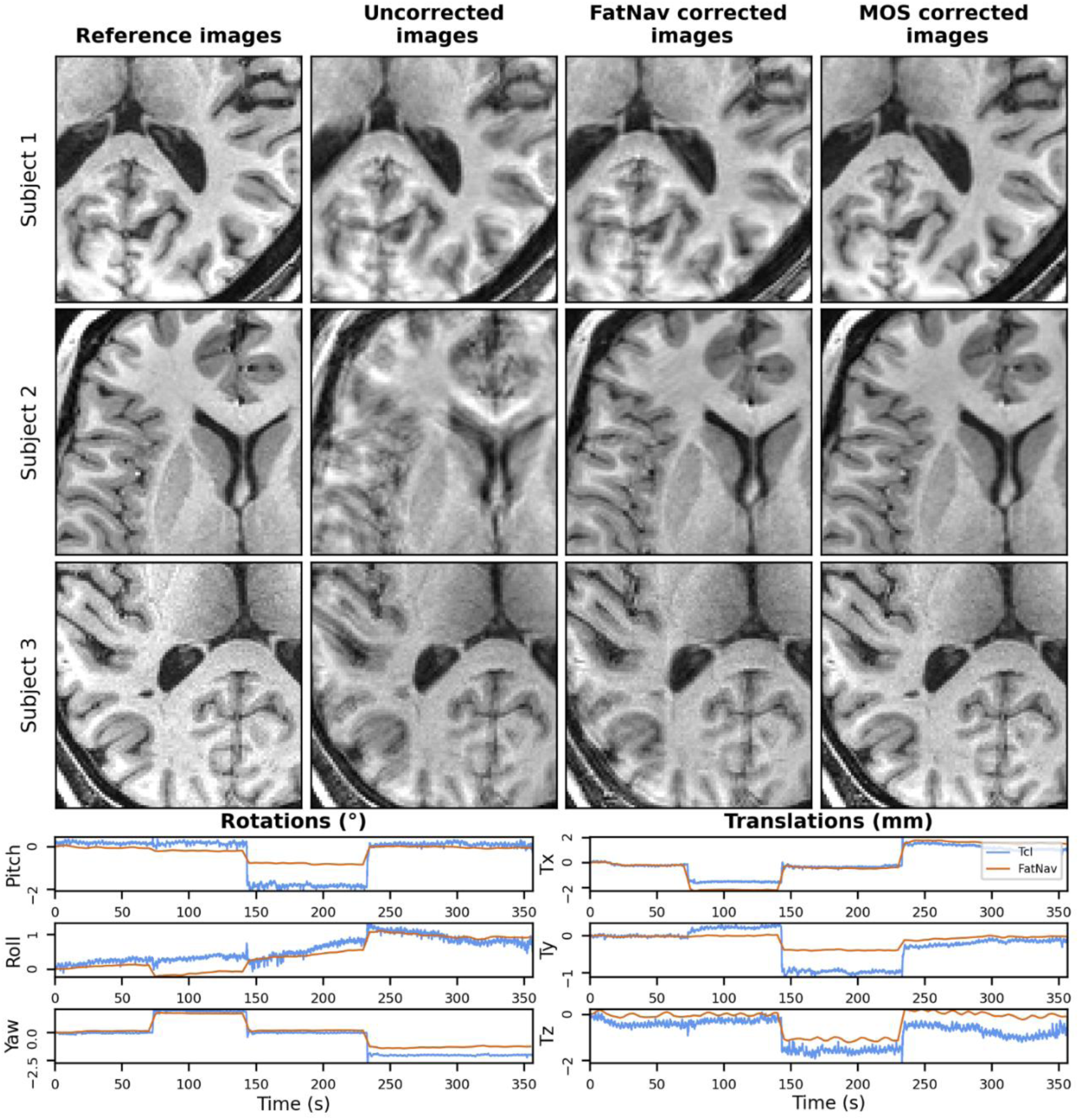
Visual difference between brain T1w-images of 3 volunteers without and with 2° head rotations, and the same images retrospectively corrected from motion by MOS and by FatNav with neck-mask. The curves show MOS and FatNav estimates of rotations and translations in X, Y and Z axes executed by the volunteer during one of the T1w-scans. Curves were baseline-corrected to zero for clarity of visualization.

In blinded image quality evaluations, three evaluators assigned significantly higher scores to MOS corrected images compared to FatNav corrected images (3.58±1.13 and 2.52±0.94, respectively; *p<0.001*; Figure 6).

**Figure 6.**
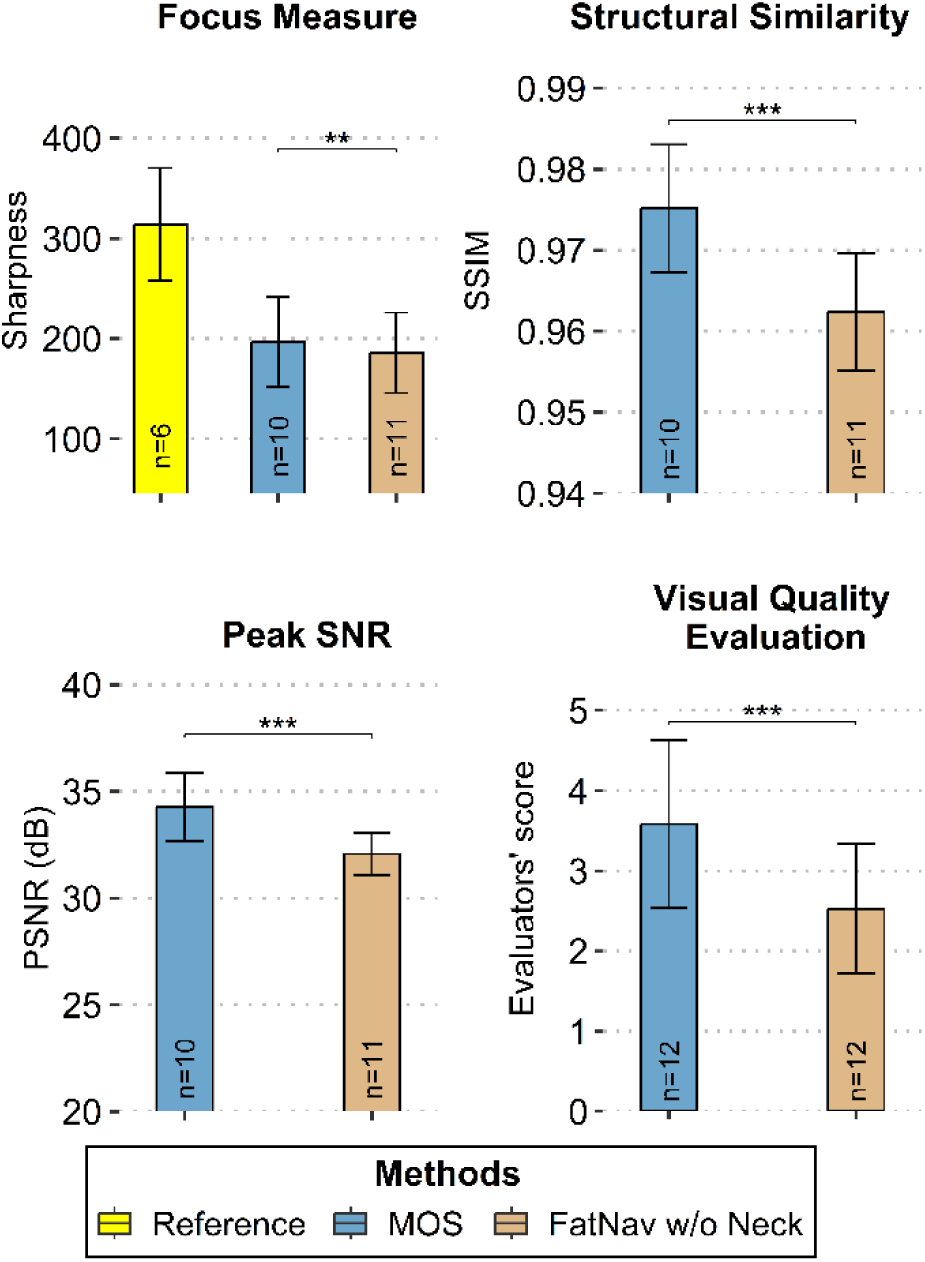
Visual and quantitative quality assessment of T1w-images collected in Experiment-2 and retrospectively corrected for motion effects using MOS and FatNav with neck-mask.

Quantitative analyses of image quality metrics further reinforced MOS’s consistent superiority in restoring T1w-image fidelity (Figure 6). SSIM comparisons between motion-free and motion-corrected images revealed higher agreement for MOS (0.98±0.008) than for FatNav (0.96±0.007; *p<0.001*). MOS also exhibited a 10.9% improvement in PSNR relative to FatNav (34.28±1.61 dB vs. 32.08±1 dB; *p*<0.001), while focus measure analyses corroborated MOS’s enhancement of image sharpness (196.8±45.33 vs. 185.81±40.22; *p*=0.009). Notably, three T1w-images were excluded from this analysis due to anomalous SSIM values, probably due to misalignment between reference images and motion-corrected images.

Finally, neither visual nor quantitative evaluations of image quality revealed statistically significant performance differences between the two FatNav variants with and without neck-masking (Supplementary data, figure 5).

## 4. Discussion

This study introduces an in-vivo approach to evaluate the accuracy of intra-MRI head-motion measurements, and consequently, the effectiveness of associated correction methodologies. The custom-developed software provided visual instruction with real-time feedback via MOS to standardize head positioning and ensuring reproducible motion across participants. Unlike other recent visual instruction systems, where subjects were instructed to follow a cross moving across the screen with their nose36, the real-time visualization of head position Is more accurate and removes the need for any calibration procedure, as both the current and target head positions are presented at the same scale on the projection screen. The head-movement guidance system exhibited low inter-subject variability (5.6%±4.3% for 2° and 2.8%±2.1% for 4°). Head translational shifts (up to 4 mm along the X-axis during yaw rotations) as well as subtle rotations around secondary non-targeted axes, consistent with natural head motion, were also observed and quantified (Supplementary data, table 1-2).

Our approach, combining visual guidance and gold-standard assessment of head movements, revealed that MOS demonstrated higher accuracy than FatNav in estimating translational motions but produced less consistent results for rotational motions. Although larger head rotations (2–4°) were more accurately captured by MOS, FatNav was more effective at detecting small-amplitude rotations around secondary axes (Figure 3).

Notably, we observe that MOS estimation errors are independent of the amplitude of head rotations (Spearman correlation R=-0.06), whereas FatNav errors increase with larger rotation amplitudes (R=0.70; p<0.001; see figure 7). To the best of our knowledge, this represents the first in vivo experimental characterization providing this level of detail. This finding is consistent with the simulation study by Andersen et al., which demonstrated that navigation errors generally increase with the magnitude of motion across various navigator types^37^. One plausible explanation is that large head rotations relative to the B₀ field result in substantial reorientation of air–tissue interfaces, leading to pronounced distortions in fat-navigator images^18^. These distortions, combined with the elastic deformation of subcutaneous fat tissue, may compromise the accuracy of FatNav, particularly during large pitch head rotations around the X-axis. Consistent with this, our experimental findings demonstrate that FatNav is less accurate in estimating translations associated with pitch rotations than with yaw rotations of the head (p=0.023).

**Figure 7.**
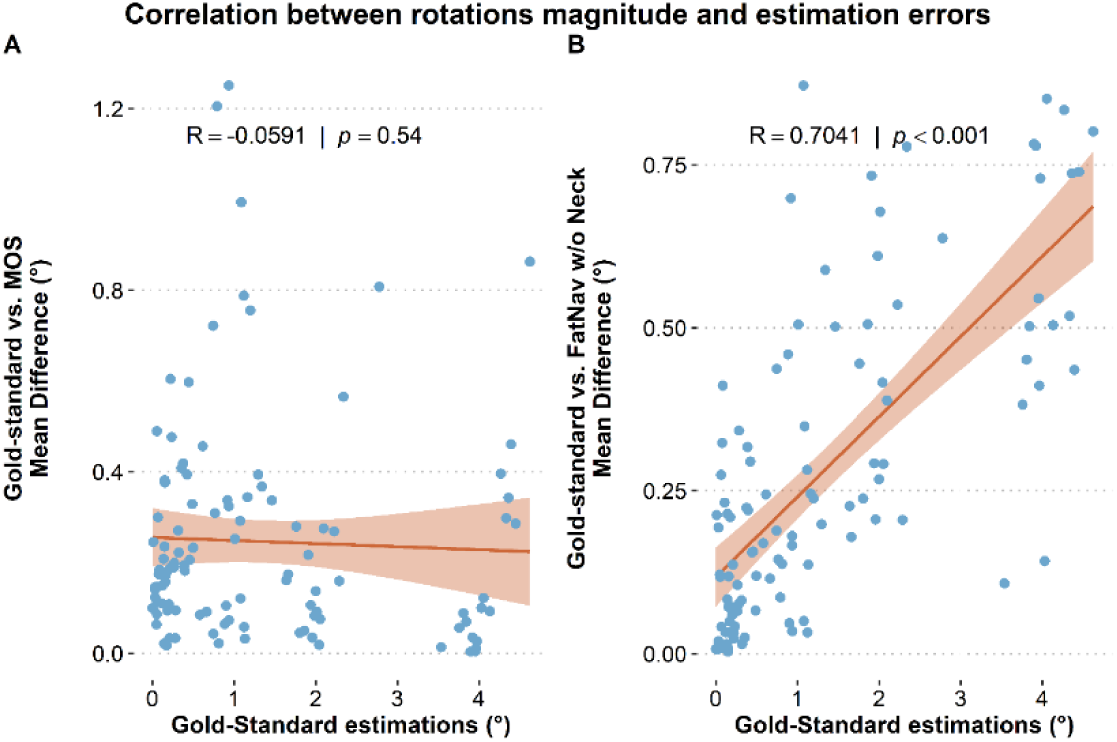
Spearman’s rank correlation between gold-standard head rotation values and the estimation errors derived from MOS and FatNav with neck-mask.

Further investigation is required to determine whether MOS shortcomings in estimating small head rotations are due to an intrinsic insensitivity to small-amplitude rotations or to confounding effects, where large rotations on one axis obscure the detection of smaller rotations on other axes. Moreover, the MOS appears to estimate yaw^+^ better than yaw^-^ rotations (Figure 3). This could be attributable to the shifting of facial features in and out of MOS’s FOV during movements. The common part of the face, useful for aligning facial surfaces over time, could therefore change according to the movement orientation and have an impact on MOS estimates.

In Experiment-2, participants moved their heads during each T1w-scan between four positions similar to those in Experiment-1. Yaw^+,^ pitch^+^ and yaw^-^ rotations were performed during each T1w-scan at 1 min 20 sec; 2 min 30 sec; and 4 minutes respectively after the start of T1w-acquisition. This choice maximizes motion-effects in the uncorrected data, while preserving the central Fourier-space sampling at pitch^+^ position. The head rotation and displacement curves, recorded during the 6-minute T1w- acquisition (Figure 5), show discrepancies between MOS and FatNav values, consistent with the errors of both systems described in Experiment-1. Marchetto et al. have reported a such discrepancies between MOS and FatNav estimates (refer to figures 4 and 5 in their study)^20^. However, due to the lack of a gold standard measurement, they were unable to resolve the performance differences between the two methods.

Quantitative and visual assessments of T1w-image quality confirmed that MOS was more effective in correcting head motion. This finding is consistent with the results of Experiment-1, as MOS exhibits greater accuracy in estimating translational head displacements as well as high-amplitude rotations, which degrade perceived image quality more severely than smaller rotations. In our study, the selected image quality metrics, applied to the skull-stripped brain region after percentile normalization, were in good agreement with expert visual assessments. Some studies have reported mismatches between image quality metrics and radiological evaluation in the presence of head motion^20^, underscoring the need for careful metric design.

Importantly, the head motion tracking evaluation approach we are proposing using a gold-standard, demonstrated sufficient sensitivity to detect improvement in FatNav estimates when a neck mask was applied to the fat-navigators. This improvement undetectable by the image quality-based approach, was most pronounced for X-axis rotations (pitch^+^) inducing elastic deformation at the head–neck junction (Figure 3-B).

So far, only Gumus et al. have conducted a comparable in-vivo assessment of head motion tracking in MRI, using SPM-registration as the gold standard. However, their analysis focused solely on unguided and primary large head rotations, which exhibited high intrasubject variability (37.5%, 8°±3°), and did not investigate the associated head translations and small rotations around the secondary axes^18^. Involuntary head motion and/or magnetic susceptibility variations from head positioning can introduce artifacts in T1w-images, potentially compromising rigid registration and thus gold-standard reliability. The next section examines these errors and their impact on gold-standard validity.

The visual instruction system with real time feedback helps reduce involuntary motion during T1w- acquisition. Although MOS motion estimates were minimal, they were used to correct T1w-images of experiment-1 for motion artifacts. To evaluate potential bias from using MOS-corrected T1w images in the comparison between MOS and FatNav, we repeated the analyses using T1w-images reconstructed without correction and with FatNav motion correction. Results show that performance differences between MOS and FatNav were consistent across all three conditions: uncorrected, MOS- corrected, and FatNav-corrected. The only exception was that MOS’s superiority in estimating translational motion during yaw^+^ rotation was no longer statistically significant without motion correction (Supplementary data, figure 6).

The distortion analyses of T1w-images previously presented demonstrate that gold-standard estimates derived from both distortion-corrected and uncorrected T1W images are highly correlated (Figure 4). Moreover, the comparison outcomes between MOS and FatNav remain largely consistent when using the distortion-corrected gold-standard values (Figure 3 and Supplementary data, figure 4). These results indicate that distortion effects are minimal within the experimental conditions of this study, and that a gold-standard based on rigid registration provides sufficiently accurate estimations for head rotations in the range of 2–4° at 3T. Nonetheless, the use of distortion-corrected gold-standard values may offer improved reliability in scenarios involving larger head movements and/or higher magnetic field strengths.

One limitation of this study is the extended acquisition time required for T1w-images at multiple head poses (4min9sec×7 ∼ 29 min). To ensure accurate gold-standard estimates via FreeSurfer registration, we prioritized high-quality 1 mm resolution images. However, in agreement with Gumus et al., who acquired 2 mm brain images every 26 seconds^18^, downsampling our 1 mm T1w-images to 2 mm and repeating the registrations did not significantly alter gold-standard motion estimates. This suggests that T1w-images with shorter acquisition times could maintain the gold-standard’s accuracy while increasing the sampling of head positions, reducing movements related to inattention, and mitigating magnetic susceptibility effects.

The in-vivo approach presented in this study enables direct quantification of errors associated with motion tracking techniques but can’t capture all facets of motion-related image correction. Notably, it does not permit assessment of how correction frequency influences the quality of reconstructed images^16^. Although we interpolated motion parameters for each Fourier-space line using the nearest available head-motion estimate, the higher temporal resolution of MOS (30 Hz vs. ∼0.4 Hz for FatNav) may confer an advantage in correcting relatively fast motions. Furthermore, this in vivo approach cannot fully account for potential estimation errors arising during rapid, large-amplitude head movements, which may distort both the fat-navigator images and the facial surface tracked by the MOS infrared camera. This limitation could be mitigated by reacquiring Fourier-space data affected by substantial motion during motion-free periods at the end of the acquisition^38,39^. To comprehensively evaluate head-motion correction strategies, it is thus recommended to combine estimation error assessment using a reliable gold standard with the evaluation based on the motion-corrected image quality.

## 5. Conclusion

In this study, we present an in-vivo framework that combines an intra-MRI head-motions visual instruction system with a rigid registration-based gold-standard to enable precise, quantitative evaluation and comparison of intra-MRI head-motion tracking techniques. This approach revealed that the markerless optical system performs best for detecting head translations and large rotations, which was corroborated by imagine quality analyses, while FatNav offers better accuracy for small rotational movements. Although image quality assessments favored the markerless optical system overall, our evaluation approach was more sensitive in detecting the subtle improvements achieved by applying neck masking to FatNav data. These findings underscore the utility of this framework for advancing and refining head-motion tracking and correction strategies.

## Acknowledgments

This work was supported by the European Research Council under the European Union’s Horizon 2020 research and innovation program (ERC Consolidator Grant 864550), LABEX CORTEX funding (ANR-11-LABX-0042) from the Université de Lyon operated by the French National Research Agency (ANR), and the National Institute of Biomedical Imaging and Bioengineering (R21EB029641 and R01EB035560).

## Data availability statement

The data and scripts that support the findings of this study are available from the corresponding author upon reasonable request.

## References

1. Gedamu EL, Gedamu A. Subject movement during multislice interleaved MR acquisitions: Prevalence and potential effect on MRI-derived brain pathology measurements and multicenter clinical trials of therapeutics for multiple sclerosis. J Magn Reson Imaging. 2012;36(2):332–343. doi:10.1002/jmri.23666

2. Sun JP, Bu CX, Dang JH, et al. Deep learning-based reconstruction enhanced image quality and lesion detection of white matter hyperintensity through in FLAIR MRI. Asian J Surg. 2025;48(1):342–349. doi:10.1016/j.asjsur.2024.09.156

3. Slipsager JM, Glimberg SL, Søgaard J, et al. Quantifying the Financial Savings of Motion Correction in Brain MRI: A Model-Based Estimate of the Costs Arising From Patient Head Motion and Potential Savings From Implementation of Motion Correction. J Magn Reson Imaging. 2020;52(3):731–738. doi:10.1002/jmri.27112

4. Andre JB, Bresnahan BW, Mossa-Basha M, et al. Toward Quantifying the Prevalence, Severity, and Cost Associated With Patient Motion During Clinical MR Examinations. J Am Coll Radiol. 2015;12(7):689–695. doi:10.1016/j.jacr.2015.03.007

5. Zong X, Nanavati S, Hung S, Li T, Lin W. Effects of motion and retrospective motion correction on the visualization and quantification of perivascular spaces in ultrahigh resolution T2-weighted images at 7T. Magn Reson Med. 2021;86(4):1944–1955. doi:10.1002/mrm.28847

6. Gal-Er B, Brackenier Y, Bonthrone AF, et al. Verifying the concordance between motion corrected and conventional MPRAGE for pediatric morphometric analysis. Published online October 2, 2024. doi:10.1101/2024.10.01.616016

7. Engström M, Mårtensson M, Avventi E, Norbeck O, Skare S. Collapsed fat navigators for brain 3D rigid body motion. Magn Reson Imaging. 2015;33(8):984–991. doi:10.1016/j.mri.2015.06.014

8. Gallichan D, Marques JP, Gruetter R. Retrospective correction of involuntary microscopic head movement using highly accelerated fat image navigators (3D FatNavs) at 7T. Magn Reson Med. 2016;75(3):1030–1039. doi:10.1002/mrm.25670

9. Laustsen M, Andersen M, Xue R, Madsen KH, Hanson LG. Tracking of rigid head motion during MRI using an EEG system. Magn Reson Med. 2022;88(2):986–1001. doi:10.1002/mrm.29251

10. Afacan O, Wallace TE, Warfield SK. Retrospective correction of head motion using measurements from an electromagnetic tracker. Magn Reson Med. 2020;83(2):427–437. doi:10.1002/mrm.27934

11. Zaitsev M, Dold C, Sakas G, Hennig J, Speck O. Magnetic resonance imaging of freely moving objects: prospective real-time motion correction using an external optical motion tracking system. NeuroImage. 2006;31(3):1038–1050. doi:10.1016/j.neuroimage.2006.01.039

12. Maclaren J, Armstrong BSR, Barrows RT, et al. Correction: Measurement and Correction of Microscopic Head Motion during Magnetic Resonance Imaging of the Brain. Hess CP, ed. PLoS ONE. 2013;8(6). doi:10.1371/annotation/a29733ae-6317-42ee-92c0-a49542e1b7c8

13. Cordero-Grande L, Ferrazzi G, Teixeira RPAG, O’Muircheartaigh J, Price AN, Hajnal JV. Motion-corrected MRI with DISORDER: Distributed and incoherent sample orders for reconstruction deblurring using encoding redundancy. Magn Reson Med. 2020;84(2):713–726. doi:10.1002/mrm.28157

14. Yoshida N, Kageyama H, Akai H, et al. Motion correction in MR image for analysis of VSRAD using generative adversarial network. Ou Y, ed. PLOS ONE. 2022;17(9):e0274576. doi:10.1371/journal.pone.0274576

15. Chatterjee S, Sciarra A, Dünnwald M, Oeltze-Jafra S, Nürnberger A, Speck O. Retrospective Motion Correction of MR Images using Prior-Assisted Deep Learning. Published online November 28, 2020. doi:10.48550/arXiv.2011.14134

16. Frost R, Wighton P, Karahanoğlu FI, et al. Markerless high-frequency prospective motion correction for neuroanatomical MRI. Magn Reson Med. 2019;82(1):126–144. doi:10.1002/mrm.27705

17. Slipsager JM, Glimberg SL, Højgaard L, et al. Comparison of prospective and retrospective motion correction in 3D-encoded neuroanatomical MRI. Magn Reson Med. 2022;87(2):629–645. doi:10.1002/mrm.28991

18. Gumus K, Keating B, White N, et al. Comparison of Optical and MR-Based Tracking. Magn Reson Med. 2015;74(3):894-902. doi:10.1002/mrm.25472

19. Bazin PL, Nijsse HE, Van Der Zwaag W, et al. Sharpness in motion corrected quantitative imaging at 7T. NeuroImage. 2020;222:117227. doi:10.1016/j.neuroimage.2020.117227

20. Marchetto E, Murphy K, Glimberg SL, Gallichan D. Robust retrospective motion correction of head motion using navigator-based and markerless motion tracking techniques. Magn Reson Med. 2023;90(4):1297–1315. doi:10.1002/mrm.29705

21. Reuter M, Tisdall MD, Qureshi A, Buckner RL, Van Der Kouwe AJW, Fischl B. Head motion during MRI acquisition reduces gray matter volume and thickness estimates. NeuroImage. 2015;107:107–115. doi:10.1016/j.neuroimage.2014.12.006

22. Avventi E, Ryden H, Norbeck O, Berglund J, Sprenger T, Skare S. Projection-based 3D/2D registration for prospective motion correction. Magn Reson Med. 2020;84(3):1534–1542. doi:10.1002/mrm.28225

23. Motion Tracking. TracInnovations. Accessed January 26, 2025. https://tracinnovations.com/motion-tracking/

24. Slipsager JM, Ellegaard AH, Glimberg SL, et al. Markerless motion tracking and correction for PET, MRI, and simultaneous PET/MRI. Zhang Q, ed. PLOS ONE. 2019;14(4):e0215524. doi:10.1371/journal.pone.0215524

25. retroMoCoBox/fatnavtools at master · dgallichan/retroMoCoBox. GitHub. Accessed January 26, 2025. https://github.com/dgallichan/retroMoCoBox/tree/master/fatnavtools

26. Griswold MA, Jakob PM, Heidemann RM, et al. Generalized autocalibrating partially parallel acquisitions (GRAPPA). Magn Reson Med. 2002;47(6):1202–1210. doi:10.1002/mrm.10171

27. Reuter M, Rosas HD, Fischl B. Highly accurate inverse consistent registration: A robust approach. NeuroImage. 2010;53(4):1181–1196. doi:10.1016/j.neuroimage.2010.07.020

28. Avants BB, Tustison NJ, Song G, Cook PA, Klein A, Gee JC. A reproducible evaluation of ANTs similarity metric performance in brain image registration. NeuroImage. 2011;54(3):2033–2044. doi:10.1016/j.neuroimage.2010.09.025

29. Smith SM. Fast robust automated brain extraction. Hum Brain Mapp. 2002;17(3):143–155. doi:10.1002/hbm.10062

30. Wang Z, Bovik AC, Sheikh HR, Simoncelli EP. Image Quality Assessment: From Error Visibility to Structural Similarity. IEEE Trans Image Process. 2004;13(4):600–612. doi:10.1109/TIP.2003.819861

31. Pertuz S, Puig D, Garcia MA. Analysis of focus measure operators for shape-from-focus. Pattern Recognit. 2013;46(5):1415–1432. doi:10.1016/j.patcog.2012.11.011

32. R: A Language and Environment for Statistical Computing. Published online 2024. https://www.R-project.org/

33. Bates D, Mächler M, Bolker B, Walker S. Fitting Linear Mixed-Effects Models Using lme4. J Stat Softw. 2015;67:1–48. doi:10.18637/jss.v067.i01

34. Fox J, Weisberg S. An R Companion to Applied Regression. Third. Sage; 2019. https://www.john-fox.ca/Companion/

35. Lenth RV. emmeans: Estimated Marginal Means, aka Least-Squares Means. Published online October 20, 2017:1.10.7. doi:10.32614/CRAN.package.emmeans

36. Berglund J, Van Niekerk A, Rydén H, et al. Prospective motion correction for diffusion weighted EPI of the brain using an optical markerless tracker. Magn Reson Med. 2021;85(3):1427–1440. doi:10.1002/mrm.28524

37. Andersen M, Laustsen M, Boer V. Accuracy investigations for volumetric head-motion navigators with and without EPI at 7 T. Magn Reson Med. 2022;88(3):1198–1211. doi:10.1002/mrm.29296

38. Tisdall MD, Hess AT, Reuter M, Meintjes EM, Fischl B, Van Der Kouwe AJW. Volumetric navigators for prospective motion correction and selective reacquisition in neuroanatomical MRI. Magn Reson Med. 2012;68(2):389–399. doi:10.1002/mrm.23228

39. Frost R, Hess AT, Okell TW, et al. Prospective motion correction and selective reacquisition using volumetric navigators for vessel-encoded arterial spin labeling dynamic angiography. Magn Reson Med. 2016;76(5):1420–1430. doi:10.1002/mrm.26040

