## Supporting materials for "An in-vivo approach to quantify intra-MRI head motion tracking accuracy: comparison of markerless optical tracking versus fat-navigators" (from another version of this preprint: version 1, 2025-04-25)

### Supplementary Materials

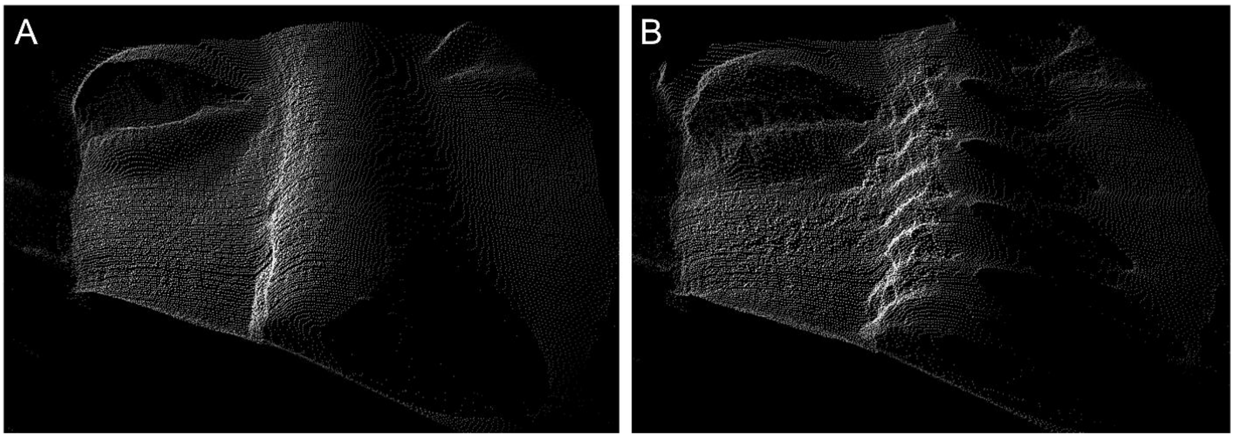


Figure S1. Point clouds of the facial surface captured by the markerless optical system through the coil's aperture in front of the right eye during a period of no movement (A) and during a period of significant head movement (B).

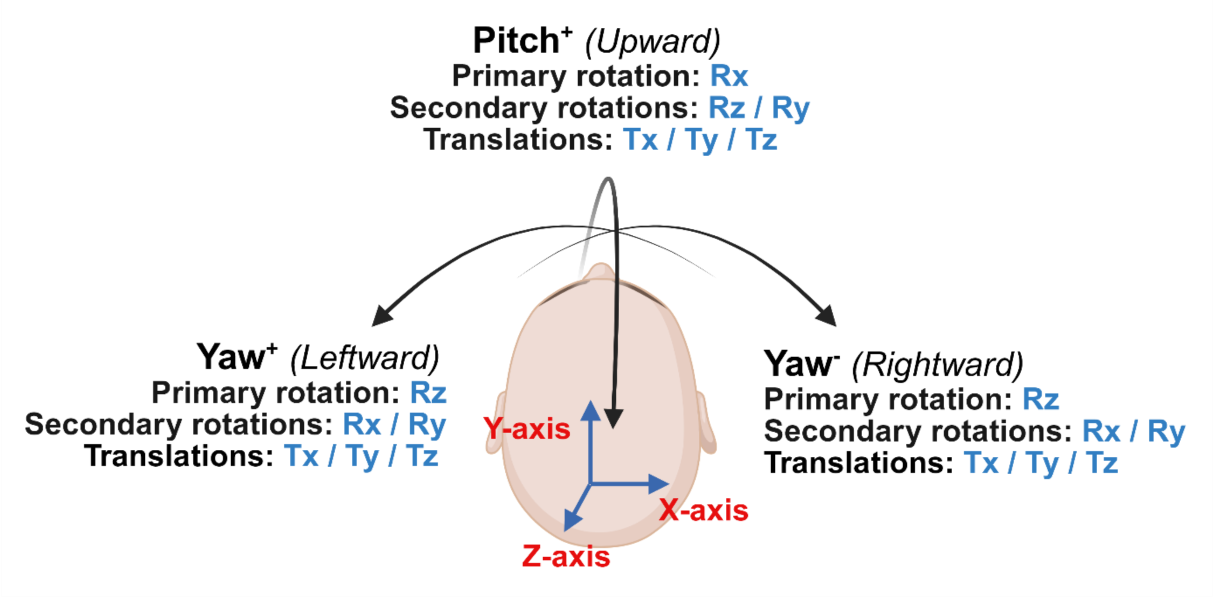


Figure S2. Primary and secondary axes of head-rotations: The primary rightward (yaw$^-$) and leftward (yaw$^+$) rotations around Z axis are associated with unintentional small secondary rotations around the Y and X axes; And the primary upward (pitch$^+$) rotation around the X axis is associated with unintentional small secondary rotations around the Y and Z axes.

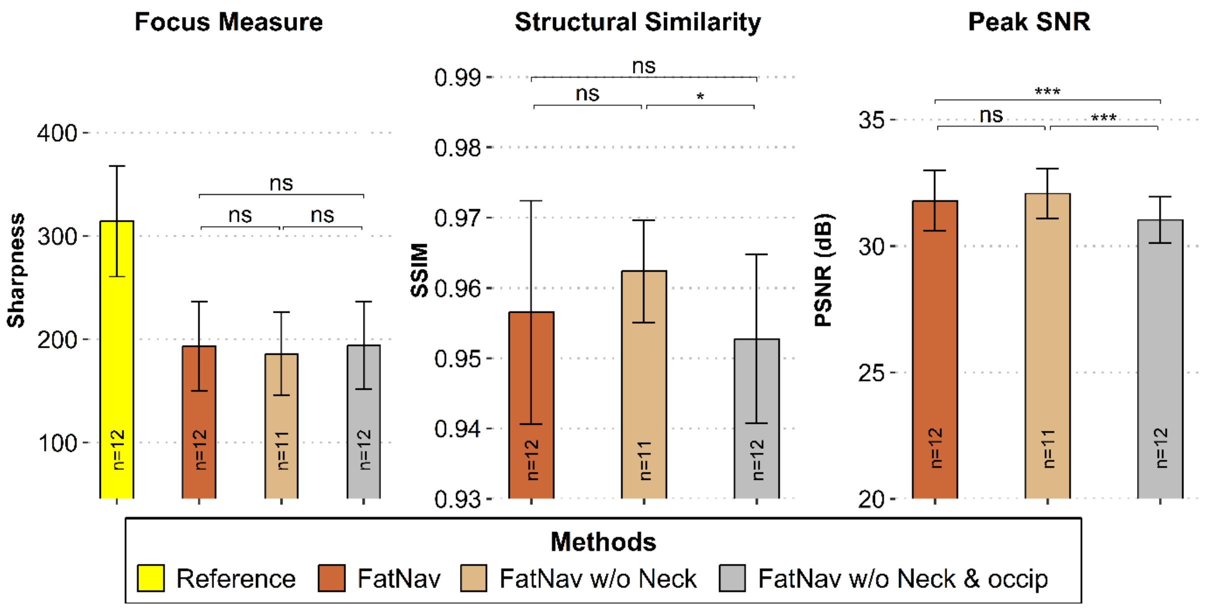


Figure S3. Quality assessment of T1-weighted images acquired in Experiment-2 and retrospectively corrected for motion effects using standard FatNav, FatNav with neck-mask and FatNav with neck and head-occipital mask.

Table S1. Mean ± SD of unintentional translation and secondary rotation estimates.

| Method | | Rx | Ry | Rz | Tx | Ty | Tz |
|---|---|---|---|---|---|---|---|
| **Gold standard** | 2° movement | **0.21±0.12** | **0.65±0.35** | **0.09±0.07** | **0.88±0.60** | **0.34±0.17** | **0.61±0.45** |
| | 4° movement | **0.22±0.20** | **0.93±0.44** | **0.19±0.16** | **1.49±1.03** | **0.52±0.40** | **0.99±0.76** |
| **MOS** | 2° movement | 0.15±0.12 | 0.71±0.46 | 0.07±0.09 | 0.96±0.72 | 0.42±0.29 | 0.83±0.47 |
| | 4° movement | 0.14±0.11 | 0.84±0.56 | 0.15±0.06 | 1.69±1.16 | 0.65±0.42 | 1.11±0.87 |
| **FatNav** | 2° movement | 0.15±0.11 | 0.58±0.33 | 0.17±0.12 | 1.29±0.89 | 0.21±0.17 | 0.67±0.59 |
| | 4° movement | 0.19±0.13 | 0.94±0.54 | 0.20±0.19 | 2.63±1.65 | 0.45±0.48 | 1.41±1.20 |
